# Design and Validation of a Low-Cost, Open-Source, 3D-Printed Otoscope

**DOI:** 10.1101/2023.08.09.23293916

**Authors:** Joshua Lowe, Hamza Bagha, Hamza Inayat, Anas Eid, Frankie Talarico, Tiffany Ni, Tarek Loubani

## Abstract

The modern otoscope is an indispensable instrument utilized by primary care physicians as the gold standard tool to diagnose an array of otologic diseases and conditions. At present, commercially available, traditional otoscopes remain cost-prohibitive to many potential users despite limited innovation since its invention in the early 19^th^ century. In this publication, the design and assembly of a low-cost, open-source, 3D-printed otoscope, the Glia Otoscope V1.0, is outlined. Subsequently, we describe the benchtop evaluation conducted, which measured several outcomes relevant to otoscopy performance against a traditional, gold standard otoscope, the Welch Allyn Rechargeable V3.5 Halogen HPX Otoscope. Measured outcomes included illuminance, correlated color temperature, color rendering index, spatial resolution, field of view, weight, battery life, and cost. Overall, the Glia Otoscope V1.0 demonstrated comparable performance across measured outcomes against the traditional otoscope. Further validation in the clinical setting is warranted as the Glia Otoscope V1.0 and its future iterations hold tremendous potential in improving access and alleviating the burden of otologic disease in lower and middle-income countries. Finally, we present a novel tool, the Otoscope Assessment Tool, which establishes a standard set of performance characteristics for benchtop evaluation of otoscope performance.

## Introduction

The otoscope is an indispensable instrument utilized by primary care physicians and health care providers.(1,2) It allows for precise and direct visualization of the outer and middle ear structures and tissues, serving as the gold standard tool for diagnosing numerous otologic diseases and conditions, including otitis media, cerumen impaction, otosclerosis, tympanic membrane perforations, infectious myringitis, and cholesteatomas.(2,3) It is most recognized for its role in diagnosing and stratifying the severity of acute otitis media (AOM), which has a global incidence rate of 709 million cases each year with 51% of cases occurring in children under 5 years of age.(4) In the United States, AOM is the most common reason for seeking medical therapy and antibiotic prescription in children under the age of 5 years, resulting in an estimated total annual health care expenditure of $2.88 billion.(4,5)

Despite its significance, especially in primary care medicine, the otoscope has largely eluded technological innovation. Its fundamental design has persisted since the late 19th century, comprising a light source, magnifying lens, and cone-shaped specula.(1,2,5) Moreover, commercially available, gold standard, traditional otoscopes (TOs), commonly used in clinical settings, remain cost-prohibitive to many potential users.(6–9) The high cost represents a significant barrier for healthcare systems and providers in low and middle-income countries (LMICs) and other low-resource settings.(6,8,9) Thus, hospitals and healthcare providers in these settings often rely on well-intentioned donations of TOs and other medical devices. However, it has been widely reported that the vast majority of donated medical devices are never actually used due to incompatibility with infrastructure or they quickly become inoperable and accumulate in a medical device “graveyard drawer” due to insurmountable logistics issues with maintenance.(10) The inadequacy of access to otoscopy in LMICs hinders healthcare providers’ abilities to effectively diagnose, classify, and treat AOM, which has resulted in a high incidence and prevalence of chronic suppurative otitis media (CSOM).(3,6) The World Health Organization (WHO) estimates that up to 330 million people globally suffer from CSOM, which is responsible for 28,000 deaths and over 2 million disability-adjusted life years annually.(3) The lack of adequate access to otoscopy has also led to the overtreatment of uncomplicated AOM and is a contributing factor toward the global antibiotic resistance crisis.(11,12) Thus, the overwhelming global burden of undiagnosed, underdiagnosed, and overtreated acute and chronic ear disease warrants greater investment into the research and development of a more accessible, low-cost otoscope that is non-inferior to gold standard devices.

Several organizations, including the WHO and Lancet have advocated for the development of more affordable medical technologies to serve those in LMICs.(8,9) This call-to-action has been heeded by various organizations around the world, including the Glia Project (Glia). Glia is an organization that develops and distributes high-quality, open-source, 3D-printed medical devices to those in LMICs, especially during periods of geopolitical instability when supply-chains are compromised.(13) In 2018, Glia validated an open-source, 3D-printed stethoscope, which cost US$1.84 to US$3.68 to produce, in a non-inferiority trial against the industry standard Littman Cardiology III.(14) Currently, over 3,000 Glia Stethoscopes have been manufactured in Canada and Gaza.(15) They are deployed in hospitals across Canada and Gaza and distributed to medical students across Canada, Kenya, and Zambia.(13–16) Glia has since developed several other open-source 3D-printed medical devices, including an otoscope called the Glia Otoscope V1.0 (GO), which is produced using accessible materials and costs US$5.00 to US$15.00 to produce. The GO aims to deliver equivalent, if not superior performance, compared to a gold standard TO.(17)

At present, only two other low-cost, open-source 3D-printed otoscope designs are available online. The first was published in 2017 in response to the 2015 Nepalese earthquake.(18) This design is a “remix” of a concept originally published on Appropedia and was further refined based on feedback from medical professionals in Nepal and the UK. However, it has not been evaluated in a clinical or research setting. The second was developed by Capobussi and Moja, who drew inspiration from the aforementioned design and created their own after refining several development elements.(19) It is the only 3D-printed otoscope design that has been studied in comparison to a commercially available TO. In their study, they compared their €5 3D-printed otoscope to a €100 Sigma F.O. LED, G.I.M.A. S.p.A. Otoscope. They reported their prototype demonstrated a similar overall quality, including the quality of vision and magnification factor, when compared to the commercially available otoscope.(19)

The main objectives of this study are threefold: (1) to describe the development and assembly process of the GO, (2) to perform a comparative benchtop evaluation of the GO against a commercially available, gold standard TO, specifically, the Welch Allyn 3.5V Rechargeable Halogen HPX Otoscope across several outcomes relevant to otoscopy performance, and (3) to present a novel tool that establishes a standard set of performance characteristics most relevant to otoscopy performance.

## Methods

### Design

The GO was originally designed by a Canadian audiologist, Frankie Talarico, through his organization, E4R Designs. He used a simplified Computer Aided Design (CAD) software named TinkerCAD and based his otoscope design on current gold standard designs of otoscopes from premium brands. The design was subsequently modified and recreated in FreeCAD, an open-source software package, in collaboration with Glia and with feedback from users in the field and public forums (Fig 1). The device is manufactured in Canada under a Health Canada Medical Device Establishment License (License #6823) to produce Class I devices.

**Fig 1.**
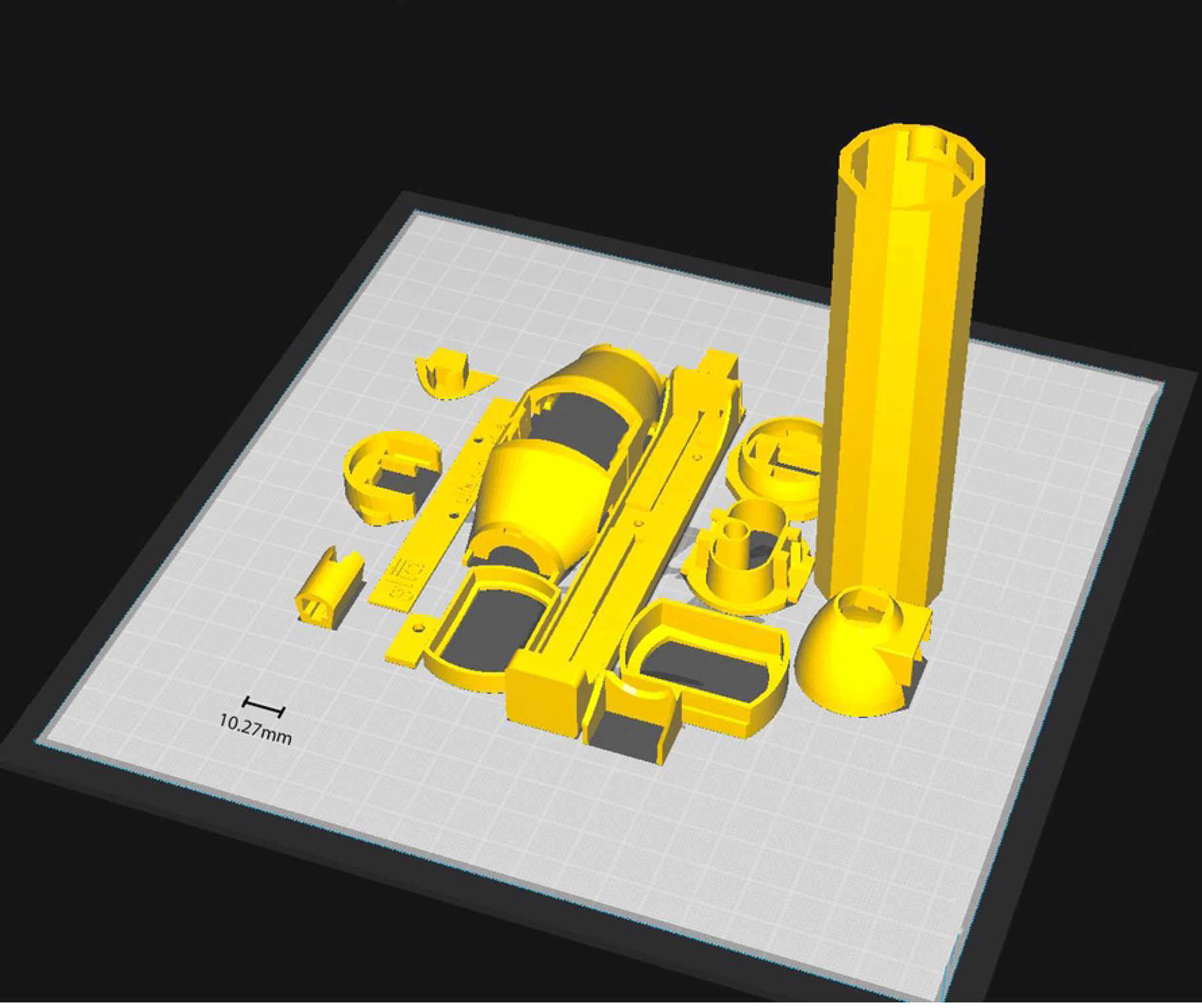
Orientation of the Glia Otoscope’s 3D-printed parts on a slicer build plate.

The 3D-printed components comprising the head and handle of the otoscope are shown in Fig 2. Additional materials required include: a AA battery holder, 10 Ohm resistor, 5mm LED light, rocker switch, 3x magnification biconvex acrylic lens, shrink tubing, and extra wiring. The GO is designed to be compatible with Welch Allyn disposable accessories, such as the specula, but can also accommodate other disposables with minor adjustments. The open-source design is freely available at https://github.com/GliaX/otoscope, [16] The fully assembled GO is displayed in Fig 3A and 3B.

**Fig 2.**
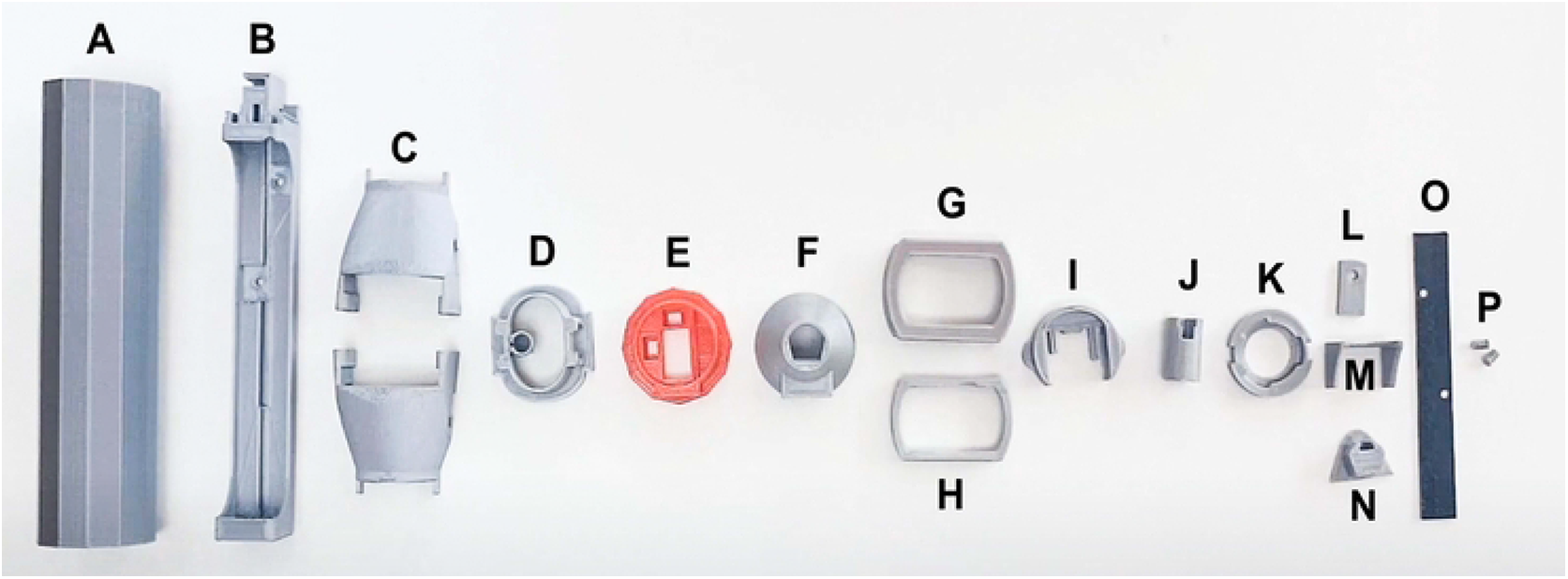
Glia Otoscope’s individual parts. A. Handle B. Battery compartment C. Two head shells D. Inner head shell E. Bottom button F. Top button G. Lens holder bottom H. Lens holder top I. Handle coupler J. Neck K. Speculum holder L. Button lock M. Button protector N. Head lock O. Name plate P. Two Name plate locks.

**Fig 3A.**
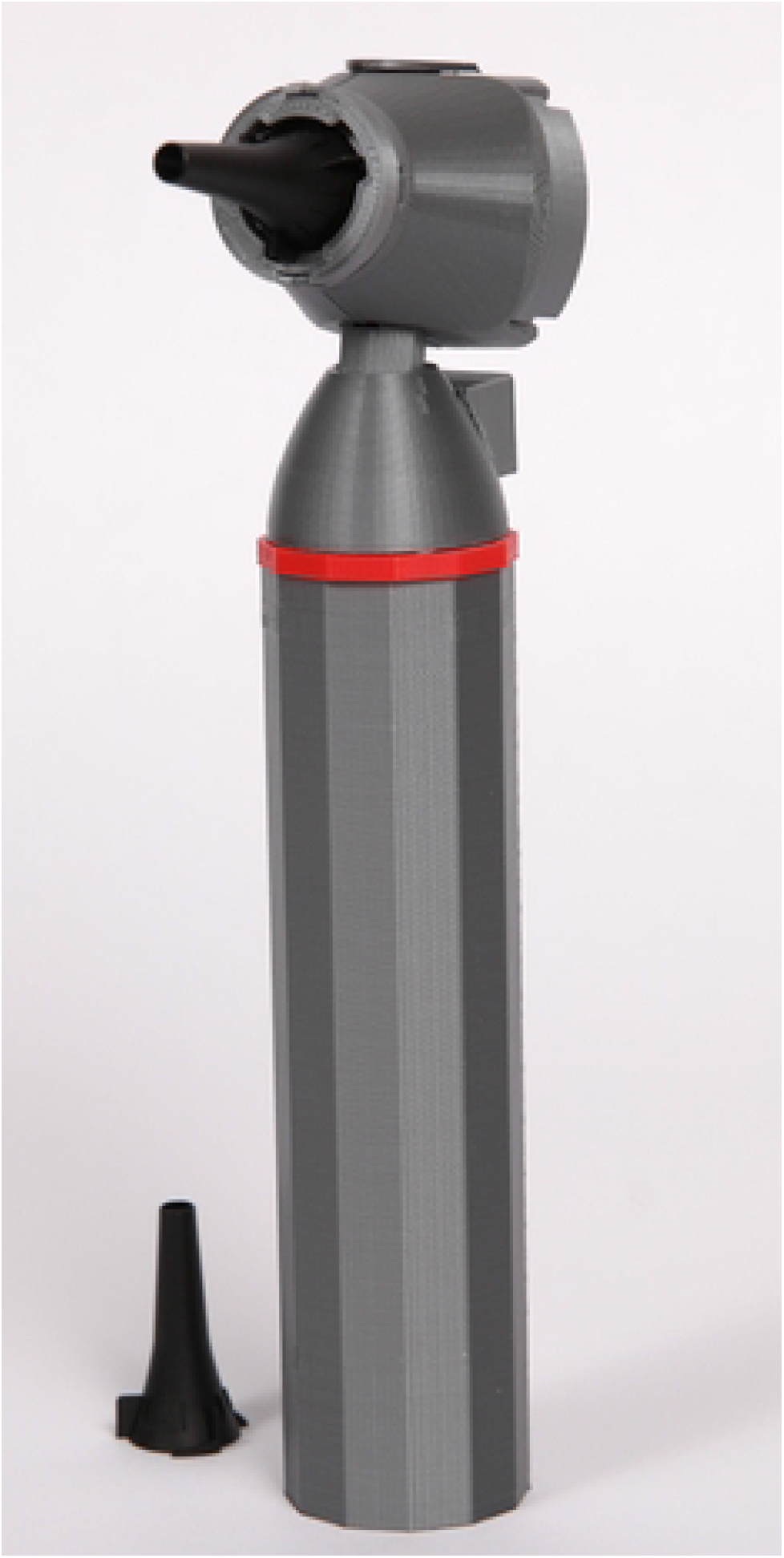
Fully assembled Glia Otoscope with Welch Allyn 4.25mm specula. Front three-fourths view.

**Fig 3B.**
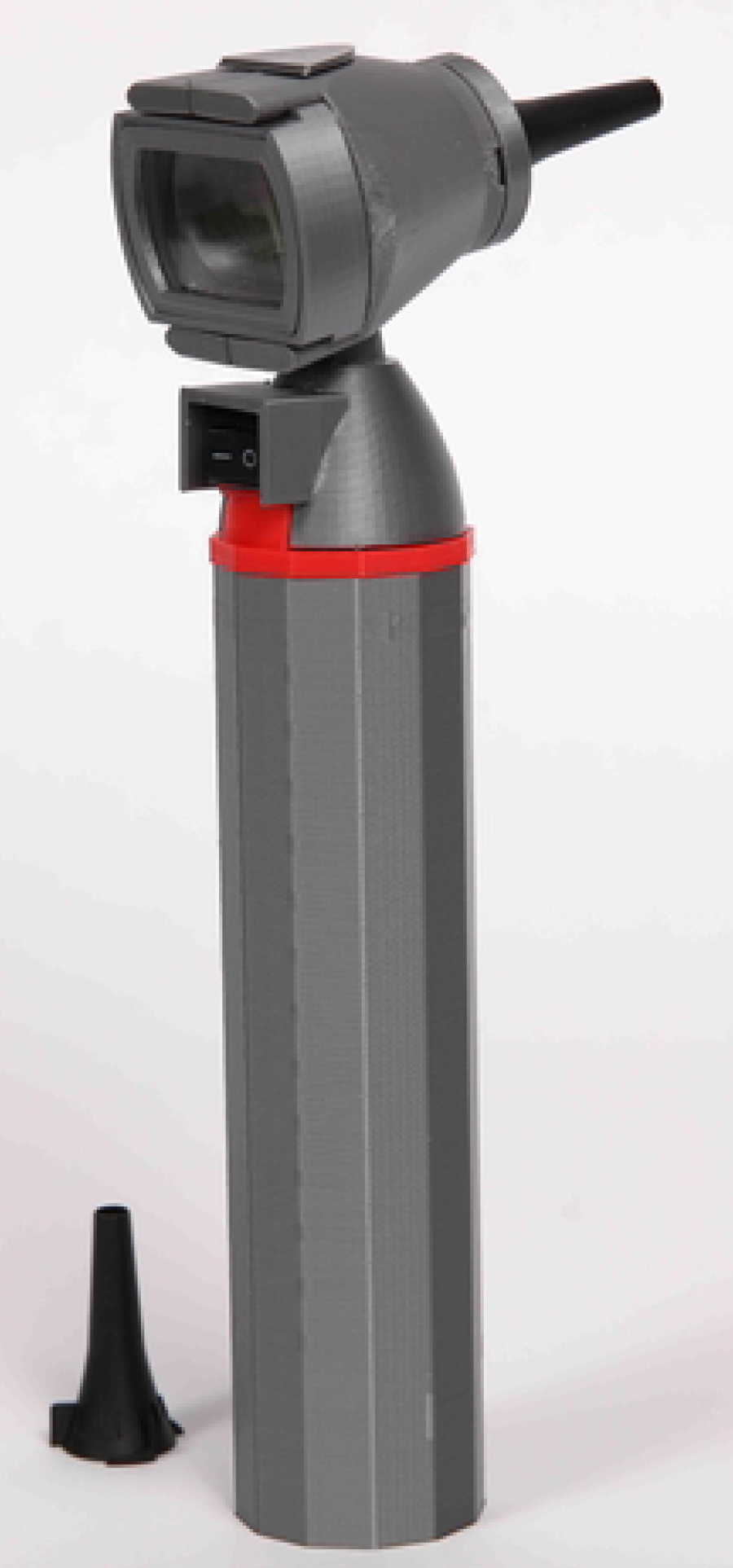
Fully assembled Glia Otoscope with Welch Allyn 4.25mm specula. Rear three-fourths view.

### 3D Printing and Assembly

All parts were printed with 1.75 mm diameter filament using a fused deposition modelling (FDM) printer. The chosen material was Polylactic Acid (PLA), and the prints were set at a 0.2mm layer height with 100% infill except for the handle, which was set at 20% infill. Assembly instructions provided by Frankie Talarico were found at https://github.com/GliaX/Otoscope and strictly adhered to.(17) Each 3D-printed part was initially dry fitted and then permanently assembled using cyanoacrylate glue.

Battery compartment: The nameplate was mounted to the back of the battery compartment using the nameplate locks. On the AA battery holder, the black wire was cut to 80mm and the red wire to 44mm. The 10-ohm resistor was soldered to the end of the red wire and an additional 130mm red wire to the other end of the resistor. The battery pack was fitted into the battery holder with the wires passing through their respective slots. The handle coupler piece was installed onto the battery holder then the bottom button piece was secured in place by the button lock.

Head: The outer head half pieces were assembled with the inner head, neck, head lock, and specula holder pieces. A 50mm red and black wire was soldered to the positive and negative terminals of the LED light, respectively, before being fitted to the inner head piece with the wires passing through their respective slots. The inner head piece was connected to the head shells, and the neck piece was attached to the head with the wires running through it. The headlock piece was glued into its slot, and lastly, the specula holder was affixed to the front of the head using cyanoacrylate glue.

Middle: The switch was connected to two black 50mm wires through soldering. The friction-fit plastic components of the switch were carefully trimmed and smoothed using a file. Subsequently, the switch was properly positioned within the button holder piece.

Otoscope Assembly: The head, middle, and battery compartment assemblies were joined together (Fig 4) while simultaneously connecting the necessary wires through soldering. The red wires from the battery and head were soldered together, and each black wire from the switch was soldered to a corresponding black wire from the battery and head. The top and bottom halves were securely bonded using cyanoacrylate glue. Two AA alkaline batteries were then inserted into the battery holder, and the handle was attached and locked into position without the use of glue.

**Fig 4.**
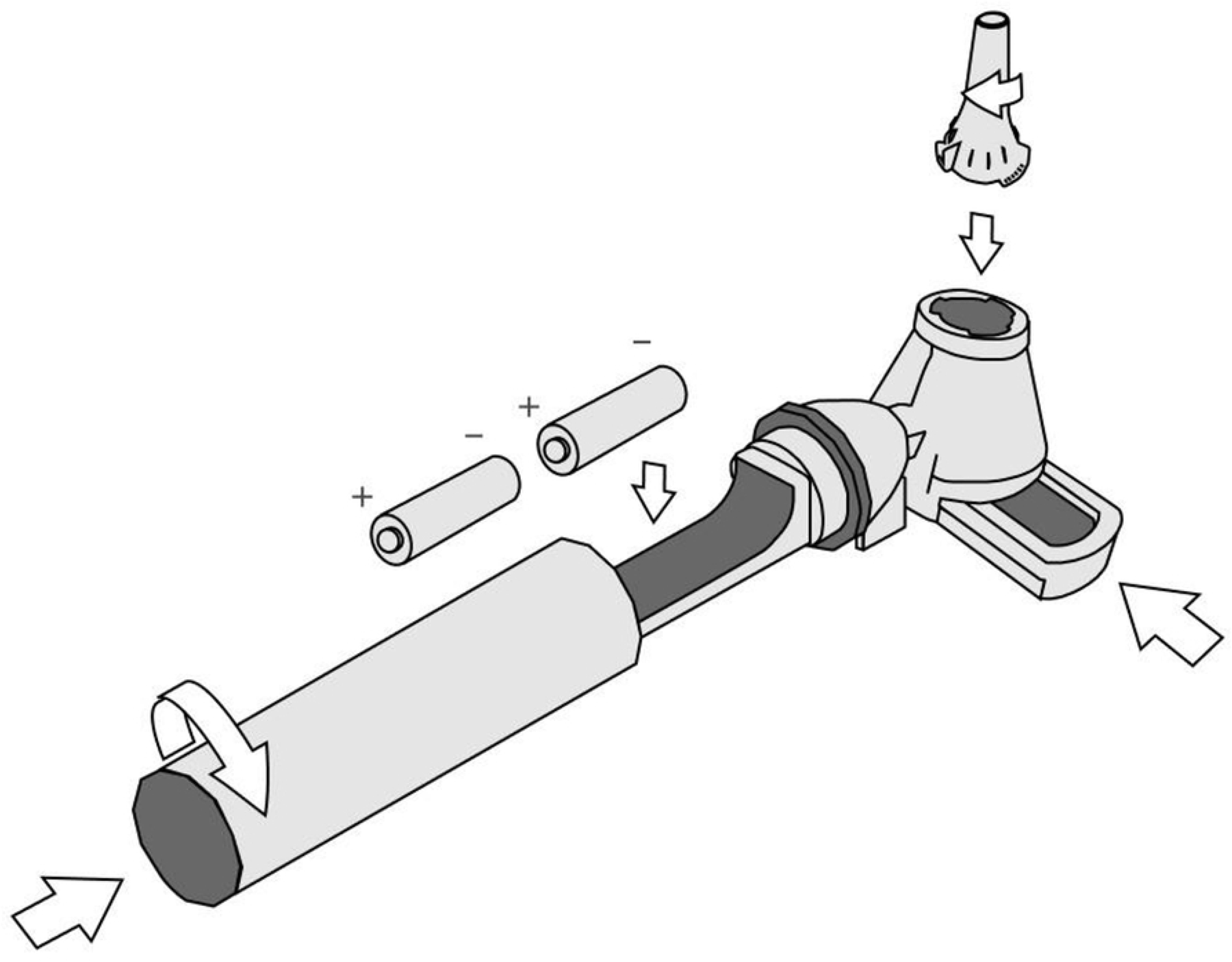
Assembly of peripheral pieces and disposables with Glia Otoscope.

Lens: The lens was sized to the lens top holder piece and cut to size with a band saw and shaped with a belt sander. It was fitted with the top and bottom lens holder pieces and installed into the otoscope assembly.

As delineated in Table 1, the cost was calculated by multiplying the weight of each 3D-printed part, determined through the Slic3r, version 1.3.0-dev, by the prevailing market price of PLA filament as of October 2022. Printer electricity expenditure was estimated using the Ontario Energy Board’s current mid-peak rates. Power usage was extrapolated over a four-hour printing period, and the resulting cost was reported in US$.

For non-3D-printed components, market prices were estimated based on our material sources listed on the Glia’s Otoscope GitHub, Bill of Materials.(17) The weight of each component and their respective costs are outlined in Table 1. This includes the AA batteries, the on/off switch, LED/resistor/wire/shrink tubing, battery holder, acrylic lens (cut to size), and the disposable specula.

### Benchtop Evaluation

The evaluation of performance characteristics was conducted by a member of the research team (JL). A Glia team member (SP) assembled a GO in London, Ontario, Canada, which was utilized for the study. A TO, specifically the Welch Allyn Rechargeable 3.5V Halogen HPX Otoscope, was loaned to the research team by the Department of Otolaryngology/Head & Neck Surgery at Queen’s University. The otoscopes are displayed side-by-side in Fig 5. All outcomes for the otoscopes were measured with a Welch Allyn KleenSpec 4.25 mm Disposable Ear Specula attached.

**Fig 5.**
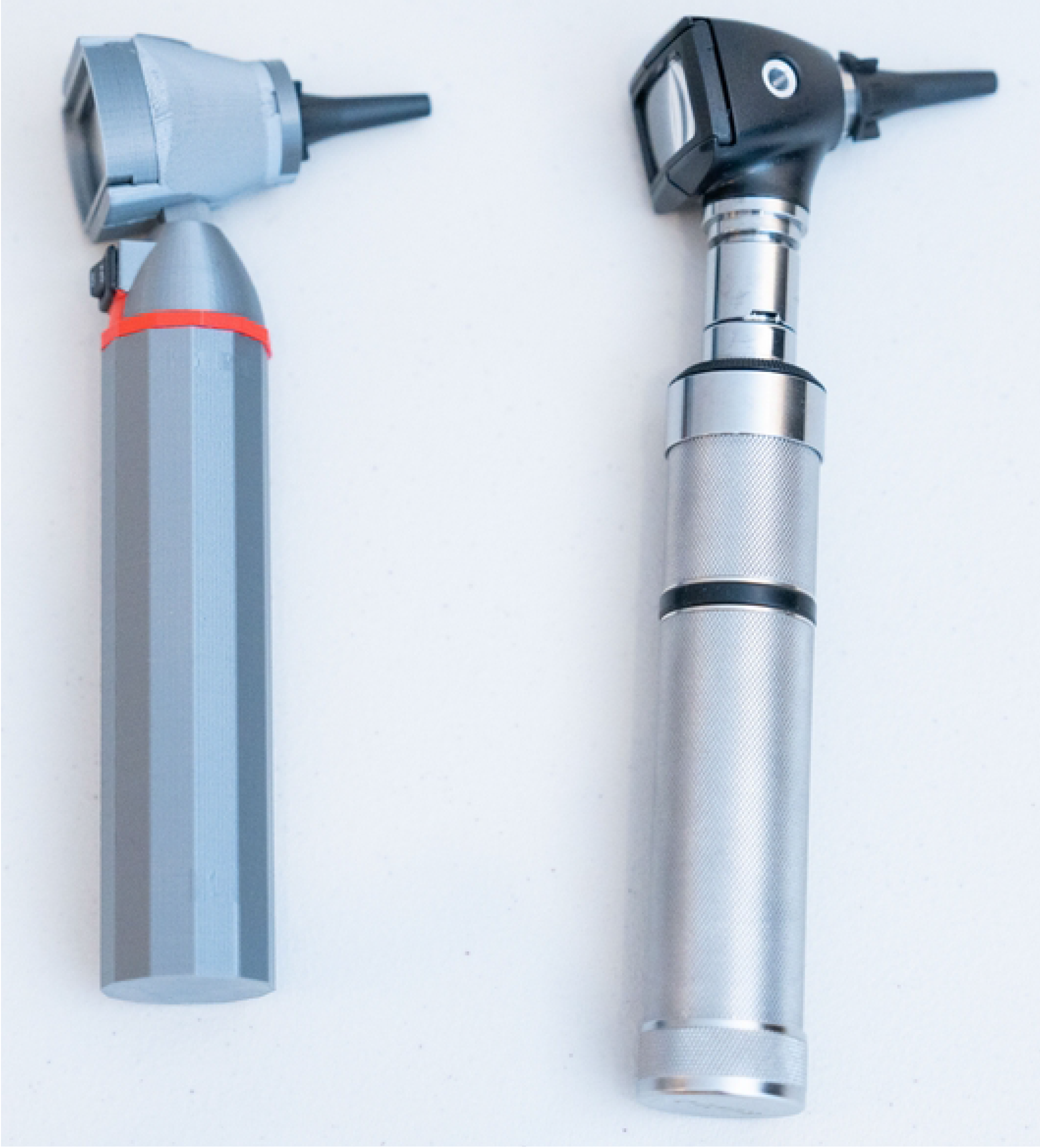
The Glia Otoscope and the Welch Allyn 3.5 V Halogen HPX Otoscope, pictured on the left and right, respectively.

At present, no standardized methodology exists to evaluate the efficacy of otoscopes. Outcomes were selected based on performance characteristics commonly reported in the literature from previously studied otoscopes, including the 3D-printed otoscope by Capobussi and Moja(19), digital otoscopes(20), smartphone otoscopes(21,22), and the solar-powered Arclight otoscope.(23) Additionally, the research team, by consensus, selected the most clinically relevant performance characteristics reported by manufacturers of gold standard TOs as outcomes.(24–26)

Illuminance, correlated color temperature (CCT), and International Commission on Illumination (CIE) R_a_ value (more generally known and herein referred to as color rendering index (CRI), were measured using the Opple Light Master III (Opple Lighting, 2022), a portable spectrometry device developed by Opple Lighting, a Dutch company with more than 23 years of experience in the professional lighting industry, operating in over 70 countries. The Opple Light Master III utilizes the AS7262 Consumer Grade Smart 6-Channel VIS Sensor, an integrated circuit (IC) that incorporates a 6-channel photodiode array.(27) This sensor samples visible wavelengths in the range of approximately 430 nm to 670 nm, with a full-width half-max of 40 nm, through an integrated aperture that provides a ±20 ° field of view. The wavelength accuracy of the CCD sensor is specified to be ±5 nm. The Opple Light Master III is highly regarded among lighting professionals for its satisfactory and comparable performance when tested against professional-grade spectrometers.(28–30) The device is capable of measuring illuminance within a range of 0-50,000 lx and color temperature within a range of 2000-250,000 K. It operates within a temperature range of -10 °C to 40 °C. The sensor of the Opple Light Master III is cited to have a deviation of approximately 5% and a resolving power of 1 lx and 1 K.(31)

The Opple Light Master III was mounted perpendicular to the surface, and a ruler was used to ensure that the tip of the specula was positioned 12.5 mm away from the sensor of the Opple Light Master III. This distance approximates half the average length of the adult human external auditory canal, an acceptable working distance for otoscopy.(32) All measurements were conducted in a completely dark environment, void of any other light source, to prevent contamination from ambient light sources. Each otoscope was powered-on to its highest brightness setting and shone towards the center of the Opple Light Master III’s sensor. The illuminance in lux (lx), correlated color temperature (CCT) in degrees of Kelvin (K), and color rendering index (CRI) of the light emitted by each otoscope were measured and recorded three times for each otoscope.

Spatial resolution was measured using the 1951 USAF resolution test chart, a widely accepted method to evaluate the spatial resolution of imaging devices in optical engineering laboratories.(33) The test chart was printed as instructed at proper scale using a Canon Pixma MP980 printer at 9600 x 2400 dpi.(34) Each otoscope was mounted such that the tip of the specula was 12.5 mm above the test chart, ensuring precise alignment. The approximate resolution limit was determined by identifying the largest element on the chart without distinct image contrast. The Group Number and Element was used to calculate the resolution with the formula shown in the following equation:

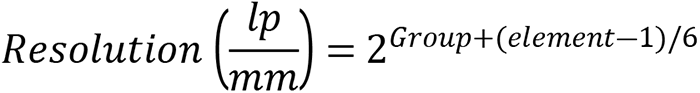

To measure the field of view (FOV), each otoscope was carefully positioned with the tip of the specula positioned 12.5 mm above the surface. Following this, a ruler was aligned within the central focus of the observed field through the otoscope, and the diameter of the field (mm) was recorded.

The weight of each otoscope was determined by positioning them individually on a pre-calibrated digital scale. Subsequently, the readings, documented in grams, were recorded. Two new 1.5 V AA alkaline batteries were placed in the GO, and the TO’s two rechargeable 3.5 V Nickel-cadmium C batteries were charged to full capacity. Both otoscopes were powered on to their maximum intensity, placed in view of a video camera, and left on. Battery life, measured in hours, was determined by calculating the duration between the first timestamp, when the otoscope was powered-on, to the second timestamp, when the otoscope failed to emit any light perceptible to the researcher (JL) on the video recording.

The Bill of Materials for the GO was tabulated by a team member (HB) and reported in Table 1. The cost of the TO was obtained from https://www.stethoscope.ca in October 2022, one of Canada’s largest authorized dealers for stethoscopes and medical equipment.(35)

**Table 1.** Bill of Materials.

| 3D-Printed Components |  |  |  |
| --- | --- | --- | --- |
| | Dimensions (mm) | Weight (g) | PLA Filament Cost (US\$) |
| Battery Holder | 18.0 X 129.5 X 18.5 | 12.3 | \$0.27 |
| Button Block | 18.5 X 13.0 X 12.7 | 1 | \$0.02 |
| Button Holder | 31.2 X 30.4 X 21.5 | 3.2 | \$0.07 |
| Button Holder Bottom | 34.0 X 34.0 X 10.0 | 4.1 | \$0.09 |
| Button Lock | 16.0 X 8.0 X 2.0 | 0.2 | \$0.00 |
| Handle Coupler | 30.5 X 22.2 X 8.0 | 0.05 | \$0.00 |
| Handle Cover | 33.0 X 33.0 X 125.5 | 30.3 | \$0.67 |
| Head Lock | 19.4 X 14.3 X 8.0 | 1.2 | \$0.03 |
| Inner Head | 34.0 X 30.8 X 13.7 | 2.1 | \$0.05 |
| Lens Holder Bottom | 40.0 X 29.1 X 10.0 | 3 | \$0.07 |
| **Lens Holder Top** | 23.7 X 36.6 X 6.0 | 1.1 | \$0.02 |
| **Name Plate Locks** | 3.0 X 9.0 X 4.5 | 0.02 | \$0.00 |
| **Name Plate** | 81.4 X 11.0 X 1.5 | 2.1 | \$0.05 |
| **Neck** | 18.0 X 10.6 X 10.0 | 1.2 | \$0.03 |
| **Outer Head Assembly** | 88.5 X 35.0 X 19.6 | 8.2 | \$0.18 |
| **Specula Holder** | 25.0 X 24.5 X 4.0 | 1.1 | \$0.02 |
| **Printer Electricity Usage (Ontario)** | 1.15 KWh Mid-Peak Usage | n/a | \$0.13 |
| **Total for 3D-Printed Components** | | **71.17** | **\$1.69** |
| **Non-3D-Printed Components** |  |  |  |
| | **Dimensions / Size**<br>**(mm)** | **Weight (g)** | **Cost (US\$)** |
| **AA Batteries (2)** | 49.7 L, D: 13.5–14.5 | 47 | \$1.57 |
| **On/Off Switch** | 10.0 X 10.0 X 15.0 | 2 | \$0.55 |
| **5 mm LED / Resistor / Wire / Shrink Tubing** | n/a | 2 | \$0.38 |
| **Battery Holder** | 107.8 X 16.2 X 13.7 | 7 | \$1.47 |
| **Acrylic Lens (cut to size)** | 22.7 X 35.6 X 5.6 | 3 | \$2.86 |
| **Disposable Specula** | 4.25 | 1 | \$0.04 |
| **Total for Non-3D Printed Components** | | 62.2 | \$6.87 |
| **TOTAL** | | **133.37 g** | **\$8.57** |

Independent, two-tailed t-tests were performed to compare mean Illuminance, CCT, and CRI. The data were analyzed using RStudio (Version 2023.6.0.421).

## Results

The measured outcomes for the GO and TO are reported in Table 2. The mean illuminance measured from the GO (M=853.33 lx, SD=109.44) was significantly lower compared to the TO (M=11486.33 lx, SD=1039.41), t(4)= -17.62, p<0.001. The measured illuminances ranged from 734 lx to 949 lx for the GO and from 10473 lx to 12550 lx for the TO. Furthermore, the light from the GO’s white LED exhibited a significantly higher mean CCT (M=9406 K, SD=853.32) compared to the TO’s halogen bulb (M=3334 K, SD=199.09), t(4)=12.00, p<0.001. The measured CCT values ranged from 8424 K to 9964 K for the GO and from 3144 K to 3541 K for the TO. Additionally, the light from the GO produced a significantly lower mean CRI (M=72.87, SD=2.71) compared to the TO (M=97.10, SD=2.17), t(4)= -12.11, p<0.001. The measured CRI values ranged from 69.9 to 75.2 for the GO and from 95.8 to 99.6 for the TO.

**Table 2.**
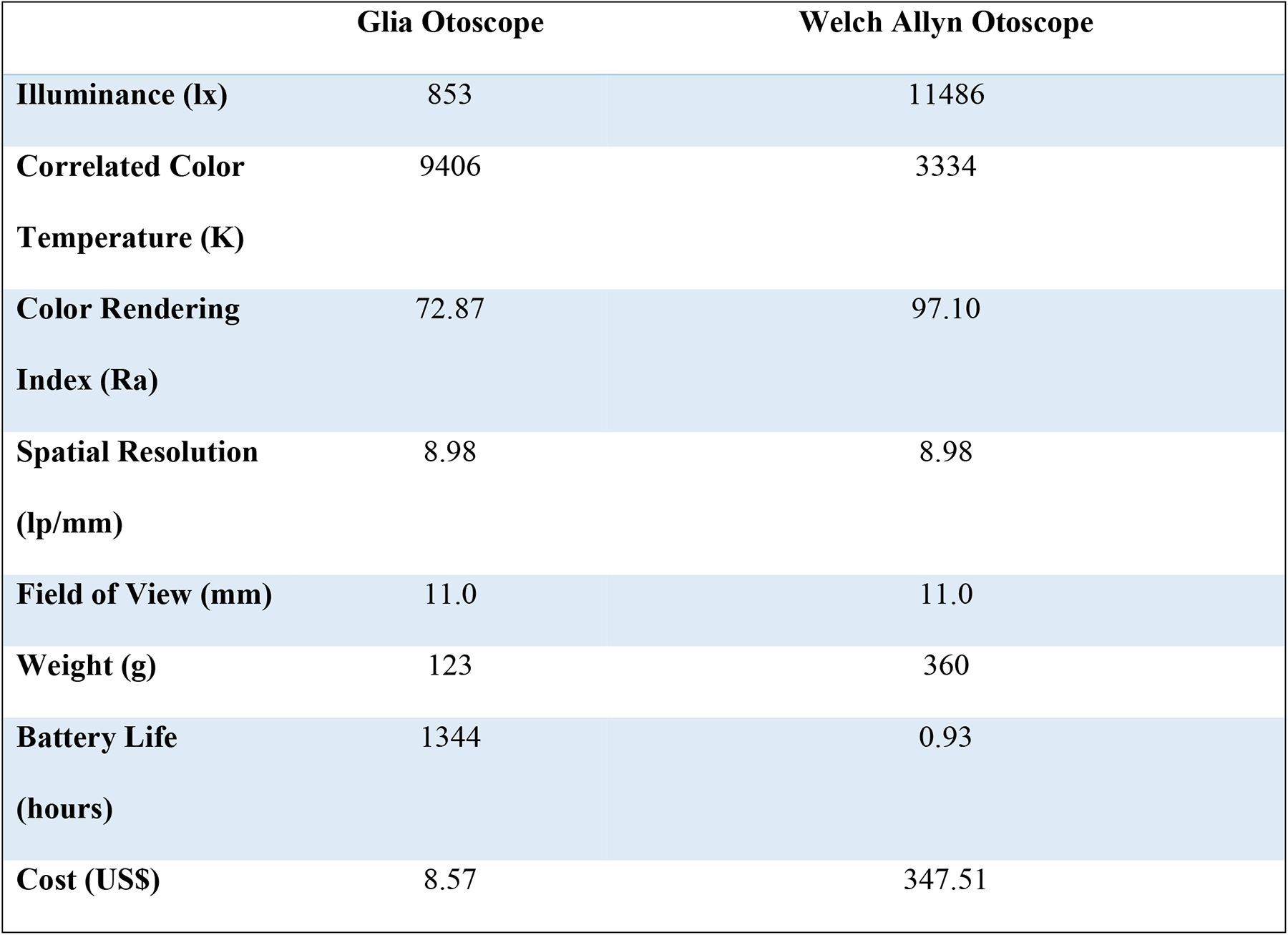
Comparison of measured outcomes between the Glia Otoscope and the Welch Allyn Rechargeable 3.5V Halogen HPX Otoscope.

The resolution and FOV were identical for the GO and TO at 8.98 lp/mm and 11.0 mm at a working distance of 12.5mm, respectively, with no variance between repeated physical measurements. The GO and TO were measured to be 123 g and 360 g, 1344 hours and 0.93 hours, and US$8.57 and US$347.51 for weight, battery life, and cost, respectively. Measurement of the GO’s battery life was discontinued after 1344 hours (8 weeks) as the light emitted was barely perceptible by the investigator (JL) and was determined to be of limited clinical utility at this point.

## Discussion

### Comparison to the gold standard, traditional otoscope

This benchtop validation study demonstrated that the Glia Otoscope (GO) is comparable to a traditional otoscope (TO), the Welch Allyn 3.5V Halogen HPX Rechargeable Otoscope, across several outcomes relevant to otoscopy performance. The GO differed from the TO in outcomes that are primarily influenced by the properties of the light source: illuminance, CCT, and CRI. The GO’s illuminance was significantly lower than the TO’s, however, according to Barriga et al.(36), who evaluated the optimal illuminance for otoscopy, its mean illuminance of 853 lx surpassed the thresholds to clearly appreciate the landmarks of the tympanic membrane and distinguish the color of the tympanic membrane well, at 215 lx and 538 lx, respectively. Additionally, it closely approached the threshold for optimal otoscopy, at 1076 lx, above which they determined there was no appreciable benefit.(36)

Second, the GO’s CCT, 9406 K, was characteristic of a cool white LED bulb producing higher emissions in the blue spectrum compared to the TO’s CCT, 3334 K, which was characteristic of a halogen bulb yielding higher emissions in the red spectrum. At present, an optimal CCT range for otoscopy has not been defined in the literature. Although CCT is a commonly reported metric in consumer materials and industry standards, its utility in evaluating otoscopy efficacy is limited, as it solely represents the color of the light emitted from the otoscope and is not indicative of operator-perceived color rendering.

Color rendering or fidelity is better represented by other spectral characteristics, such as CRI, which is a quantitative measure of the ability of an illuminant to reveal the colors of various objects faithfully in comparison with an ideal or natural light source.(37) A minimum standard CRI for otoscopy has not been defined in the literature, however, various Canadian government-regulated sectors, including Food Inspection and Museology and Conservation, recommend a minimum acceptable limit of 70, considered “fair”, for general lighting and applications, and the GO would meet this threshold with a CRI of 72.87.(38,39) For healthcare and other industries that perform intricate tasks where high color fidelity is critical, the minimum acceptable limit for CRI ranges from 80 to 90, which is considered “good to excellent”, and the TO would meet this threshold with a CRI of 97.10.(38,40) Yet, it is unclear whether this discrepancy in CRI values would result in any clinically significant difference in diagnostic acuity. For example, erythema of the tympanic membrane has been demonstrated to be nonspecific and less important than position and mobility for diagnosing AOM.(41) It is conceivable the GO’s CRI would be adequate for otoscopy and would yield equivalent clinical outcomes to the TO as the diagnosis of otologic pathology factors-in a myriad of signs and symptoms, often with greater likelihood ratios and predictive values than the subtleties of colors on otoscopy.(41) Notably, in recent years, the CIE, the international authority for developing standards in the fields of light and lighting, has expressed the necessity to update the CRI as a measure because LED light sources have frequently demonstrated significant disagreement between CRI and overall perceived color rendering.(42) Therefore, the measured CRI of the GO may not represent its color rendering ability accurately. Several alternative standards have been proposed, including CIE-Rf, Color Quality Scale, and Gamut Area Scale, however, they require further validation and have not yet succeeded in replacing CRI as the standard for color rendering.(43–45)

In summary, the GO demonstrated exceptional performance for its cost across lighting performance characteristics and it is likely capable of non-inferiority compared to the TO in the clinical setting. Future iterations can be readily and inexpensively enhanced by improving the light source, including sourcing a higher quality LED or increasing the number of LEDs to the design. Future validation studies should exclude CCT as a primary outcome as its utility in otoscopy performance is limited, and alternatives to CRI Ra, such as CIE-Rf, should be strongly considered.

The GO and TO demonstrated equivalent performance in spatial resolution and FOV, outcomes governed by their lens properties, indicating that their numerical apertures and resolving capabilities were also equivalent. Consequently, the 3x magnification, injection-molded acrylic bi-convex lens employed in the GO, sourced at US$2.86 (Table 1), represents a comparable, cost-effective solution.

Finally, the GO exhibited superior performance in outcomes determined by the materials employed: weight, battery life, and cost. The GO weighed approximately one-third of the TO’s mass due to the lower density of the 3D printing filament used in the GO compared to the metal and plastic used in the TO. This reduction in may afford its user enhanced control and maneuverability during otoscopy, which would reduce the risk of irritation, injury, and infection.(46) Next, the GO’s battery life outperformed that of the TO by a considerable margin. The disparity is multifactorial and is implicated by the intrinsic differences in efficiencies between the LED and halogen light sources as well as the non-rechargeable and rechargeable batteries. Moreover, the GO lacked a voltage regulator or power management circuit, and thus it continued to operate at progressively lower voltages, gradually dimming until it was almost indiscernible, whereas the TO likely incorporated a voltage regulator to ensure it only operated above a certain voltage to maintain a specified illuminance. Thus, implementation of a voltage regulator should be considered in future iterations of the GO and future validation studies should plot a decay curve to better account for the decrease in illuminance with battery depletion and degradation. Of note, the TO was not new and had been utilized in clinic for an unspecified period, which suggests battery degradation was a factor; however, the measured battery life of 0.93 hours does not deviate substantially from the manufacturer’s specified duration of 1 hour.(7) Regardless, the longevity of battery life demonstrated by the GO holds remarkable promise that would be particularly advantageous in LMICs, where electrical infrastructure is often unreliable and compromised.(8) Finally, the cost of the GO was over 40 times lower than that of the TO. Of note, labor costs were not factored in for the cost of the GO as it is freely accessible as an open-source project and individuals can personally assemble it if they so choose to. Assembly time for individuals building their own GO was reported to be approximately 3 hours initially for the average user then gradually reduced to as low as 30 minutes with subsequent builds as experience and familiarity were gained. Regardless, the wide cost disparity highlights the tremendous potential for cost savings that can be realized by employing the GO and other 3D-printed medical devices in LMICs. Overall, the GO demonstrated comparable performance in parameters associated with the materials used compared to the TO.

In summary, this benchtop validation study demonstrates that the GO, at a cost of US$8.57, delivers comparable performance to the Welch Allyn 3.5 V Halogen HPX across several outcomes relevant to otoscopy performance. Thus, it is a viable low-cost alternative for healthcare workers in LMICs and low-resource settings who are unable to access or afford a commercially available TO.

### Comparison to a previously studied open-source, 3D-printed otoscope

The GO and the open-source, 3D-printed otoscope developed by Capobussi and Moja (19) share a few similarities. First, both otoscopes adhere to the conventional structure and configuration of a TO, comprising of a cone-shaped head outfitted with a light source and lens mounted on a handle housing the power source. Additionally, the quantity of 3D-printed elements and non-3D-printed components needed for assembly is comparable. Specifically, the GO requires 15 3D-printed parts and 9 non-3D-printed components (Table 1). In contrast, Capobussi and Moja otoscope requires 9 3D-printed parts—with one prototype using FDM and the second prototype using stereolithography (SLA)—and 11 non-3D-printed components. These counts include batteries and specula as non-3D-printed parts. Second, the cost of the 3D printing filament and the electricity used to operate the 3D printer were comparable at US$1.67 for the GO and US$1.92 for Capobussi and Moja’s otoscope. Finally, the overall assembly time for individuals is comparable with an estimated initial assembly time of 3 hours for the average user with it significantly decreasing to as low as 30 minutes with increased experience and familiarity.

There are also a few important differences between the GO and Capobussi and Moja’s otoscope. First, the position and mounting of the LED lights are substantially different. The GO’s LED is positioned in a custom mount within the head, which allows for unobstructed light emission directly through the specula aperture, whereas the Capobussi and Moja otoscope arranges 6 LEDs in a “ring shape around the visual pathway” and emits its light directly through a translucent resin before the specula aperture.

Second, the type of lens incorporated in each otoscope is distinct despite both providing 3x magnification. The GO employs a bi-convex acrylic lens whereas the Capobussi and Moja otoscope employs a Fresnel lens. The bi-convex acrylic lens confers a few important advantages due to its intrinsic physical properties, including superior optical resolution and increased durability (47); however, it is also more costly and difficult to process, which could limit its accessibility in certain settings. An acrylic Fresnel lens was initially prototyped in the GO, and is likely a reasonable substitute in resource-limited settings, however, there were concerns regarding the degree of optical distortion and decreased durability, which could depreciate performance to an substandard level.(47) Furthermore, incorporating the bi-convex acrylic lens did not result in a substantial increase in the GO’s final cost, did not markedly affect processing time, and it is a common commodity that can be accessed globally.

Finally, the otoscopes differed in reported outcomes: illuminance, CCT, and FOV. Of note, it is not possible to draw any definitive conclusions due to variations in methodologies, and therefore, only general comparisons can be made. The GO exhibited a higher illuminance of 853 lx, in contrast to the Capobussi and Moja otoscope, which reported 70 lx. This result was unanticipated given the GO utilized a single 5 mm, 3.3 V LED bulb, while Capobussi and Moja design incorporated six 5mm, 3.3 V LED bulbs. Capobussi and Moja did not specify the capacity or type of AA batteries utilized nor the configuration of the circuit. It is possible their LEDs did not operate at their full capacity if their voltage or current requirements were not met. In their study, Capobussi and Moja stated they “…used a professional exposimeter (Bowens flash meter III, Sekonic Electronics Inc, Japan) and converted in lux in order to account for the distance between light source and target”. Since illuminance, expressed in lux units, is independent of distance, it is plausible that they were conveying luminous intensity in units of candelas, which is contingent upon the distance from the light source to the surface. It is also possible that their working distance substantially exceeded the otoscopy standard or their design may not have transmitted light as effectively given its transmission of light through the translucent resin of the head. The GO’s illuminance proved to be more akin to the values presented in the product sheets of commercially available otoscopes, such as the Welch Allyn Pocket LED and Pocket Plus LED Otoscope, which reported 1240 lx and 1540 lx, respectively, at a 50 mm working distance with a 4 mm specula (25). Moreover, the GO emitted a cooler light, 9406 K, compared to their otoscope, 6700 K, however, as stated previously, CCT has limited utility in the evaluation of otoscopy efficacy.

Finally, the FOV of the GO was noticeably larger than the FOV of Capobussi and Moja’s otoscope, with diameters of 11.0 mm and 4 mm, respectively. Their methodology was not specified, however, it is conceivable they reported the aperture diameter of the specula or otoscope head rather than the field diameter as viewed through the lens as 4 mm would be suboptimal for otoscopy, given the average adult tympanic membrane approximates 8 mm to 10 mm in diameter.(48) Thus, there were key differences identified between the GO and Capobussi and Moja’s otoscope, including position of the LEDs, the type of lens, and certain measured outcomes.

In summary, several similarities and differences were identified between the open-source, 3D-printed otoscopes. Comparable performance between the otoscopes is plausible if they were optimized and subject to the same methodology, considering their equivalence in power source and magnification. Importantly, Capobussi and Moja highlighted the modularity of their design and the ability of the consumer-maker community to rapidly iterate and improve their designs.

They substantiated this claim by rapidly designing and producing an otoscope with an UV light source. Moreover, they proposed additional innovations, such as enhancing its magnification by integrating a second Fresnel lens, as well as expanding its versatility by designing a handle capable of accommodating interchangeable heads for otoscopy, dermoscopy, and ophthalmoscopy.(19) The open-source design of both 3D-printed otoscopes affords significant potential and fostering the consumer-maker community will help further accelerate innovation in this field. Overall, both otoscopes demonstrate the opportunity for open-source, 3D-printed medical devices to improve access and positively impact the health and well-being of patients globally.

### Standardized outcomes for the benchtop evaluation of otoscope performance

The Otoscope Assessment Tool (OAT) establishes a standardized set of outcomes most relevant to otoscopy performance (S1). The tool evaluates a test otoscope against a traditional under three categories of performance characteristics: light source, lens, and miscellaneous. Working distance for light source and lens characteristics is set at 12.5mm. For the light source, measured outcomes include illuminance (lx) and Colour Rendering Index (CIE-Rf). For the lens, measured outcomes include spatial resolution (lp/mm), and field of view (mm). Finally, Miscellaneous outcomes include weight (g), battery life (hours), and cost ($USD).

### Limitations

Several limitations were noted in the present study. One primary constraint was the limited sample size. Initially, this was considered adequate for a preliminary benchtop study, accompanied by basic statistical analysis. Logistical challenges and resource limitations during the COVID-19 pandemic further justified this approach. Future studies requiring more complex analyses could increase the sample size of each otoscope to account for intra-model variances.

Next, although the Opple Light Master III is highly regarded among lighting professionals and enthusiasts as a cost-effective solution to spectrophotometry, it is not a true spectrophotometer. It samples visible wavelengths through its 6-channel photodiode array then transmits the signal to the IC, which uses an algorithm to compare the input against a set of reference values. It approximates the true value with an accuracy cited to be within 5% and resolving powers of 1 lx and 1 K. Thus, the level of accuracy and precision of the outcomes measured with the Opple Light Master III, illuminance, CCT, and CRI, were likely diminished to some degree compared to those potentially attained with a true spectrophotometer.

Finally, the GO’s open-source design introduces variability depending on sourcing of materials, printing, and assembly, which may result in variable performance of a device assembled outside Glia when compared to our measured outcomes. Indeed, variability appears inherent in the open-source 3D printing landscape and represents a potential limitation to the external validity of the GO. Capobussi and Moja asserted that even a millimetric difference in LED positioning for their device led to suboptimal performance.(19) Glia’s otoscope is designed to be simple to assemble and test, thereby reducing this variability. In addition to the Bill of Materials, Glia provides an assembly manual, lens creation guide, and assembly video. Since its release in 2018, the Glia Otoscope V1.0 has been assembled by professional and amateur makers with excellent results, so there is some evidence that consistency can be achieved with the current design and instruction sets. Nevertheless, device construction by other groups and individuals may result in differences, especially if those groups do not faithfully observe the Bill of Materials and instructions.

### Future research

As of May 2023, considerable progress has been made on the development of the GO V2.0. Its design has been fully remodeled in FreeCAD, an open-source software, and simplified so that it requires fewer 3D-printed parts and a minimal use of adhesives, facilitating easier assembly and improved durability. The complete set of FreeCAD models for all Glia Otoscope V2.0 components are freely available at https://github.com/GliaX/Otoscope. After the Glia Otoscope V2.0’s design and production are finalized, a benchtop non-inferiority trial is warranted to evaluate its performance against the GO V1.0 and a TO using the standards specified by the Performance of Otoscopy Evaluation Tool. If the GO V2.0 demonstrates non-inferiority compared to the TO, a follow-up clinical study would be warranted, in which intra and inter-rater reliability in the diagnosis of otologic pathology could be compared. An adjacent qualitative evaluation could be conducted by administering a Likert-scale questionnaire that surveys operator preferences and ease of use.

Another area of future research that warrants investigation is the GO’s utility in medical education. Otoscopy training has been identified as an area of deficiency in medical students with low overall satisfaction and confidence regarding their exposure and ability to diagnose pathology.(49) In clinical practice, otolaryngology-related disorders constitute 20-40% of all family medicine encounters and greater than 50% of pediatric primary care patients present with otolaryngology-related complaints.(50) Considering 49.44% of Canadian medical students matched to either Family Medicine or Pediatrics in the 2023 Canadian residency match (51), greater investment of resources towards improving competency in otoscopy is imperative. The GO could be distributed to medical students, at relatively low-cost, which would improve access and provide more opportunities to practice otoscopy. Subsequently, diagnostic acuity and confidence in assessing otologic pathology could be evaluated.

Finally, multidisciplinary research is paramount to ensure proper implementation and scale-up of the GO in LMICs and low-resource settings. Specifically, innovations in processes will be critical to the real-world adoption of the GO. Best practices for establishing product development partnerships, managing cultural barriers, and implementing international product standards must be investigated further. Additionally, direct engagement and dialogue with healthcare professionals and key stakeholders in LMICs and low-resource settings will be essential for ensuring feasibility and comprehensive understanding of cost, wider economic effects, challenges of distribution, human resources, and energy infrastructure. Ultimately, innovations in processes will be required to ensure the GO is not just accessible but also acceptable in meeting the unique needs of those across various LMICs and low-resource settings.(8)

## Data Availability

All Results files are available at https://github.com/GliaX/Otoscope.

## Acknowledgements

We would like to thank Carrie Wakem, Dr. Melanie Columbus, Jennifer Wilson, Steve Plimmer, Dr. Hugh Kim, Dr. Mohammed Chamma, and the Department of Otolaryngology/Head & Neck Surgery at Queen’s University for their support of this project and its authors.

## Supporting Information

S1 Fig. The Otoscope Assessment Tool (OAT) for the technical evaluation of clinically relevant outcomes for 3D-printed otoscope.

## References

1. Kravetz RE. A look back. The Otoscope. Am J Gastroenterol. 2002 Feb;97(2):470–470.

2. Mankowski NL, Raggio BS. Otoscope Exam. In: StatPearls [Internet]. Treasure Island (FL): StatPearls Publishing; 2023 [cited 2023 May 23]. Available from: http://www.ncbi.nlm.nih.gov/books/NBK553163/

3. World Health Organization. Chronic suppurative otitis media : burden of illness and management options [Internet]. World Health Organization; 2004 [cited 2023 Jun 6]. Available from: https://apps.who.int/iris/handle/10665/42941

4. Monasta L, Ronfani L, Marchetti F, Montico M, Brumatti LV, Bavcar A, et al. Burden of Disease Caused by Otitis Media: Systematic Review and Global Estimates. PLOS ONE. 2012 Apr 30;7(4):e36226.

5. Ahmed S, Shapiro NL, Bhattacharyya N. Incremental health care utilization and costs for acute otitis media in children. The Laryngoscope. 2014;124(1):301–5.

6. Blaikie A, Sandford-Smith J, Tuteja SY, Williams CD, O’Callaghan C. Arclight: a pocket ophthalmoscope for the 21st century. BMJ. 2016 Dec 14;355:i6637.

7. stethoscopeca [Internet]. [cited 2023 Jun 7]. Welch Allyn 3.5 V Halogen HPX Otoscope Set with MacroView Otoscope wit. Available from: https://stethoscope.ca/products/welch-allyn-3-5-v-halogen-hpx-otoscope-set-with-macroview-otoscope-with-throat-illuminator

8. Howitt P, Darzi A, Yang GZ, Ashrafian H, Atun R, Barlow J, et al. Technologies for global health. The Lancet. 2012 Aug;380(9840):507–35.

9. World Health Organization. WHO compendium of innovative health technologies for low-resource settings 2016-2017 [Internet]. 2018. Available from: https://www.who.int/medical_devices/innovation/compendium/en/

10. Marks IH, Thomas H, Bakhet M, Fitzgerald E. Medical equipment donation in low-resource settings: a review of the literature and guidelines for surgery and anaesthesia in low-income and middle-income countries. BMJ Glob Health. 2019 Sep 29;4(5):e001785.

11. Choosing Wisely Canada [Internet]. [cited 2023 Jun 7]. Emergency Medicine. Available from: https://choosingwiselycanada.org/recommendation/emergency-medicine/

12. Respiratory Tract Infections – Antibiotic Prescribing: Prescribing of Antibiotics for Self-Limiting Respiratory Tract Infections in Adults and Children in Primary Care [Internet]. London: National Institute for Health and Clinical Excellence (NICE); 2008 [cited 2023 May 29]. (National Institute for Health and Care Excellence: Guidelines). Available from: http://www.ncbi.nlm.nih.gov/books/NBK53632/

13. Researchers develop 3D-printed stethoscope for use in war zones, low-income areas | CTV News [Internet]. [cited 2023 Jun 7]. Available from: https://www.ctvnews.ca/health/researchers-develop-3d-printed-stethoscope-for-use-in-war-zones-low-income-areas-1.3842994

14. Pavlosky A, Glauche J, Chambers S, Al-Alawi M, Yanev K, Loubani T. Validation of an effective, low cost, Free/open access 3D-printed stethoscope. PLOS ONE. 2018 Mar 14;13(3):e0193087.

15. The Glia Project [@Glia_Intl]. 472 #3dprinted stethoscopes packaged up and ready to send to first year #medstudents @SchulichMedDent, @uOttawaMed and @uoftmedicine! All #opensource and all with #equalcare in mind. https://t.co/JE9JlDyiFU [Internet]. Twitter. 2022 [cited 2023 Jun 7]. Available from: https://twitter.com/Glia_Intl/status/1494764289898561547

16. Glia [Internet]. [cited 2023 Jun 7]. The Glia Stethoscope Project. Available from: https://glia.org/pages/stethoscope

17. Glia [Internet]. [cited 2023 Jun 7]. The Glia Otoscope Project. Available from: https://glia.org/pages/otoscope

18. 3D printed otoscope – Appropedia [Internet]. [cited 2023 Jun 7]. Available from: https://www.appropedia.org/3D_printed_otoscope

19. Capobussi M, Moja L. An open-access and inexpensive 3D printed otoscope for low-resource settings and health crises. 3D Print Med. 2021 Nov 17;7(1):36.

20. Tötterman M, Jukarainen S, Sinkkonen ST, Klockars T. A Comparison of Four Digital Otoscopes in a Teleconsultation Setting. The Laryngoscope. 2020;130(6):1572–6.

21. Chan KN, Silverstein A, Bryan LN, McCracken CE, Little WK, Shane AL. Comparison of a Smartphone Otoscope and Conventional Otoscope in the Diagnosis and Management of Acute Otitis Media. Clin Pediatr (Phila). 2019 Mar 1;58(3):302–6.

22. Kravchychyn FDB, Meurer AT de O, Nogueira MHSDP, Balieiro FO, Balsalobre F de A, Barauna Filho IS, et al. Smartphone-enabled otoscopy: method evaluation in clinical practice. Braz J Otorhinolaryngol. 2023 Jan 1;89(1):122–7.

23. Hey SY, Buckley JC, Shahsavari S, Kousha O, Haddow KA, Blaikie A, et al. A mixed methods comparative evaluation of a low cost otoscope (Arclight) with a traditional device in twenty-one clinicians. Clin Otolaryngol. 2019 Nov;44(6):1101–4.

24. Dispomed [Internet]. [cited 2023 Jun 7]. Welch Allyn MacroView Otoscope. Available from: https://www.dispomed.com/products/welch-allyn-macroview-otoscope/

25. Welch Allyn Pocket PLUS LED Otoscope 22880 (BLACK) with Soft carry case : Amazon.in: Industrial & Scientific [Internet]. [cited 2023 Jun 7]. Available from: https://www.amazon.in/OTICA-Welch-Pocket-LED-Otoscope/dp/B07YFLCP5N?th=1

26. Axiom Medical Supplies [Internet]. [cited 2023 Jun 7]. Welch Allyn Otoscope Diagnostic Type 2.5 Volt LED Pocket – M-1010733-4466 | Each. Available from: https://axiommedicals.com/products/welch-allyn-otoscope-diagnostic-type-2-5-volt-led-pocket-m-1010733-4466-each

27. ams [Internet]. [cited 2023 Apr 10]. AS7262 Spectral Sensing Engine. Available from: https://ams.com/en/as7262

28. https://www.facebook.com/1lumenflashlights. https://1lumen.com/. 2022 [cited 2023 Jun 7]. Cheap device to measure color temperature of light / flashlights (Opple Light Master 3 PRO). Available from: https://1lumen.com/gear-review/opple-light-master-3-pro/

29. Opple Light Master III (G3) discussion thread (Cheap device for measuring Lux, CCT + CRI) – Flashlight Modding and DIY Parts – BudgetLightForum.com [Internet]. [cited 2023 Apr 10]. Available from: https://budgetlightforum.com/t/opple-light-master-iii-g3-discussion-thread-cheap-device-for-measuring-lux-cct-cri/67890/471

30. Opple Light Master Pro 3 – Light at Speed [Internet]. 2022 [cited 2023 Jun 7]. Available from: https://www.youtube.com/watch?v=onara4C-hQc

31. OPPLE Lighting | Light-master-III [Internet]. [cited 2023 Jun 7]. Available from: https://www.opple.eu/en/product/indoor/light-master/light-master-g3/light-master-iii

32. Bedard N, Shope T, Hoberman A, Haralam MA, Shaikh N, Kovačević J, et al. Light field otoscope design for 3D in vivo imaging of the middle ear. Biomed Opt Express. 2016 Dec 14;8(1):260–72.

33. Pfefer J, Agrawal A. A review of consensus test methods for established medical imaging modalities and their implications for optical coherence tomography. In: Design and Quality for Biomedical Technologies V [Internet]. SPIE; 2012 [cited 2023 May 30]. p. 65–74. Available from: https://www.spiedigitallibrary.org/conference-proceedings-of-spie/8215/82150D/A-review-of-consensus-test-methods-for-established-medical-imaging/10.1117/12.912371.full

34. Lens Rating System Used in These Pages [Internet]. [cited 2023 Jun 7]. Available from: http://www.takinami.com/yoshihiko/photo/lens_test/pdml-procedure_c.html

35. Stethoscope.ca (@stethoscope_ca) • Instagram photos and videos [Internet]. [cited 2023 Jun 7]. Available from: https://www.instagram.com/stethoscope_ca/

36. Barriga F, Schwartz RH, Hayden GF. Adequate Illumination for Otoscopy: Variations due to Power Source, Bulb, and Head and Speculum Design. Am J Dis Child. 1986 Dec 1;140(12):1237–40.

37. color rendering index (CRI) (of a light source) – Illuminating Engineering Society [Internet]. [cited 2023 Jun 7]. Available from: https://www.ies.org/definitions/color-rendering-index-cri-of-a-light-source/

38. Government of Canada CFIA. Lighting in an establishment [Internet]. 2017 [cited 2023 May 20]. Available from: https://inspection.canada.ca/preventive-controls/lighting/eng/1511197522669/1528205027203

39. Institute CC. Light, ultraviolet and infrared [Internet]. 2017 [cited 2023 May 22]. Available from: https://www.canada.ca/en/conservation-institute/services/agents-deterioration/light.html

40. Soltic S, Chalmers A. Optimization of LED Lighting for Clinical Settings. J Healthc Eng. 2019 Aug 27;2019:5016013.

41. Rothman R, Owens T, Simel DL. Does This Child Have Acute Otitis Media? JAMA. 2003 Sep 24;290(12):1633–40.

42. Position Statement on CRI and Colour Quality Metrics (October 15, 2015) | CIE [Internet]. [cited 2023 May 22]. Available from: https://cie.co.at/publications/position-statement-cri-and-colour-quality-metrics-october-15-2015

43. Jost S, Cauwerts C, Avouac P. CIE 2017 color fidelity index Rf: a better index to predict perceived color difference? J Opt Soc Am A Opt Image Sci Vis. 2018 Apr 1;35(4):B202–13.

44. Davis W, Ohno Y. Color quality scale. Opt Eng. 2010 Mar;49(3):033602.

45. Lee S, Yoon HC. LED lighting system for better color rendition space: the effect of color rendering index. J Asian Archit Build Eng. 2021 Sep 3;20(5):556–65.

46. Ear Examination | HealthLink BC [Internet]. [cited 2023 Jun 7]. Available from: https://www.healthlinkbc.ca/tests-treatments-medications/medical-tests/ear-examination

47. Davis A, Kühnlenz F. Optical Design using Fresnel Lenses. Opt Photonik. 2007;2(4):52–5.

48. Jeught SV der, Dirckx JJJ. Real-time structured light-based otoscopy for quantitative measurement of eardrum deformation. J Biomed Opt. 2017 Jan;22(1):016008.

49. Niermeyer WL, Philips RHW, Essig GF, Moberly AC. Diagnostic accuracy and confidence for otoscopy: Are medical students receiving sufficient training? The Laryngoscope. 2019 Aug;129(8):1891–7.

50. Campisi P, Asaria J, Brown D. Undergraduate Otolaryngology Education in Canadian Medical Schools. The Laryngoscope. 2008;118(11):1941–50.

51. CaRMS Forum Presentation is now live! – CaRMS [Internet]. [cited 2023 Jun 7]. Available from: https://www.carms.ca/news/carms-forum-presentation-is-now-live-3-2/

